# Humoral and T-Cell Responses Following MVA-BN Booster Vaccination Against Mpox Virus Clades Ib and IIb

**DOI:** 10.1101/2025.07.07.25330986

**Authors:** Mazzotta Valentina, Matusali Giulia, Cimini Eleonora, Caioli Alessandro, Esvan Rozenn, Colavita Francesca, Tartaglia Eleonora, Paulicelli Jessica, Micheli Giulia, Bettini Aurora, Notari Stefania, Giacinta Alessandro, Bordi Licia, Gili Simona, Siddu Andrea, Girardi Enrico, Maggi Fabrizio, Antinori Andrea

**Affiliations:** Regional AIDS Reference Centre, National Institute for Infectious Diseases, Lazzaro Spallanzani – IRCCS, Rome, Italy; Laboratory of Virology and Laboratories of Biosecurity, National Institute for Infectious Diseases, Lazzaro Spallanzani -IRCCS, Rome, Italy; Laboratory of Cellular Immunology and Pharmacology, National Institute for Infectious Diseases, Lazzaro Spallanzani -IRCCS, Rome, Italy; Department of Epidemiology, National Institute for Infectious Diseases, Lazzaro Spallanzani – IRCCS, Rome, Italy; Unit of Health Promotion and Prevention, Directorate of Health and Integration, Lazio Region, Rome, Italy; Scientific Direction, National Institute for Infectious Diseases, Lazzaro Spallanzani -IRCCS, Rome, Italy; Health Direction, National Institute for Infectious Diseases, Lazzaro Spallanzani -IRCCS, Rome, Italy

## Abstract

The immunity conferred by MVA-BN vaccination against mpox appears to wane over time, particularly in individuals without prior smallpox vaccination, highlighting the potential benefit of a booster dose. We evaluated both humoral and cellular immune responses elicited by an MVA-BN booster administered at least 2 years after the primary course in 37 individuals at risk for mpox. Our results show that the booster restores and enhances both neutralising antibodies and T-cell responses, supporting the immunological value of booster strategies for individuals at ongoing risk of mpox. Moreover, we observed significant increases in IgG titres against both MPXV clades IIb and Ib, suggesting cross-protection among *Orthopoxviruses*. These findings support the use of a booster dose two years after the MVA-BN primary cycle to strengthen immunity in high-risk populations. They may inform future vaccination guidelines and public health recommendations in the evolving epidemiological context.

**Short Abstract:** We assessed humoral and cellular immune responses following an MVA-BN booster administered ≥2 years after the primary course in individuals at risk for mpox. The booster significantly increased IgG titers against MPXV (clades Ib and IIb), neutralising antibodies and T-cell responses, suggesting the booster’s benefit.

## MAIN TEXT

The mpox virus (MPXV) continues to circulate worldwide^1^. The emergence of Clade Ib in Africa, followed by imported cases in non-endemic countries, and the ongoing transmission of Clade IIb worldwide, led the WHO to renew the Public Health Emergency of International Concern^2^. Despite the widespread use of MVA-BN vaccination, its long-term protection remains uncertain. Several studies suggest that immunity wanes over time, especially in individuals without a history of smallpox vaccination^3 4^, highlighting the potential usefulness of a booster dose^5 6^. Previously, Ilchmann et al. investigated the humoral response to an MVA-BN booster using a vaccinia virus-based assay, showing a rapid and robust VACV-neutralising antibody (nAb) response after the booster^7^, but no data are available on the specific antibody response against MPXV (let alone against different clades) or on the T-cell response after the booster dose. In this scenario, booster administration remains controversial^8^ and it is recommended only by a few countries^9^.

We assessed both humoral and T-cell responses elicited by an MVA-BN booster dose, also characterising IgG response against different MPXV clades (Ib and IIb).

### Methods

We included people not historically vaccinated for smallpox, who received a primary two-dose course of MVA-BN vaccination at least two years before and who accepted the administration of a booster dose, as per Italian Ministry of Health guidelines^10 11^. All subjects eligible signed a written informed consent to be enrolled in the study. The Mpox-Vac study was approved by the INMI Lazzaro Spallanzani Ethical Committee (approval number 41z, Register of Non-COVID Trials 2022). Demographic and behavioural characteristics linked to mpox exposure and information regarding HIV status, CD4 count, HIV pre-exposure prophylaxis (PrEP) uptake and any history of previous STIs were collected. For evaluation of booster-elicited immunity, participants were sampled on the day of the booster (baseline, T1) and one month later (T2). We measured anti-MPXV IgG antibody titres using an indirect immunofluorescence assay (IFA) based on in-house prepared slides with Vero E6 cells infected with either clade IIb MPXV or a clade Ib MPXV. MPXV (Clade IIb) nAb titres were measured using the 50% plaque reduction neutralisation test (PRNT_50_)

The Interferon-γ-producing T-cell-specific response to the MVA-BN vaccine was analysed by ELISpot assay (Detailed laboratory methods in supplements).

Antibody titres and T-cell-specific responses to the MVA-BN vaccine were compared using the Wilcoxon paired test to assess levels before and after vaccination. Fisher’s exact test was used to compare the frequency of reactive samples before and after the booster dose. Analyses were performed using GraphPad Prism version 10 (GraphPad Software) for Windows statistical software; p□<□0.05 was considered statistically significant.

### Results

Among the 37 participants included, all were male at birth, 95% identifying themselves as cisgender men who have sex with men. Median age was 39 years (34-46), and 5 of them were people with HIV (PWH). Table 1 summarises the characteristics of the study population.

**Table 1.** Characteristics of the study population.

| Characteristic | N = 37<sup>1</sup> |
| --- | --- |
| Sexual orientation |  |
| Bisexual | 1 (2.7%) |
| Transgender | 1 (2.7%) |
| MSM | 35 (95%) |
| Age | 39.9 (34.5, 46.3) |
| PrEP use |  |
| No | 8 (22%) |
| Yes | 3 (8.1%) |
| Not reported | 26 (70%) |
| >=1 STI over previous year | 7 (19%) |
| PLWH | 5 (14%) |
| PLWH on effective ART | 5 (100%) |
| CD4 cell count, cells/mm3 |  |
| <200 | 0 (0%) |
| 200-500 | 2 (40%) |
| >500 | 3 (60%) |
| **Route of administration of first dose** |  |
| Intradermal | 33 (89%) |
| Subcutaneous | 4 (11%) |
| Subcutaneous | 0 (0%) |
| **Route of administration of booster dose** |  |
| Intradermal | 4 (11%) |
| Subcutaneous | 33 (89%) |
| Time between second dose and booster dose, months | 25.70 (25.03, 26.58) |

<sup>1</sup>n (%); Median (Q1, Q3)

One month after booster vaccination, IgG titres significantly increased from baseline for both clades Ib and IIb (p < 0.0001), and the proportion of reactive individuals reached 100% in both groups (p < 0.0001). However, a difference of less than one dilution was observed between the two clade groups (p=0.0148) (Figure 1A-B). Likewise, nAb titres against clade IIb significantly increased from baseline to one month after the booster (p<0.0001), with the reactivity rate rising from 8% to 76% (p<0.0001) (Figure 1C). Similarly, one month after the booster dose, the MVA-BN T-specific response was higher than before vaccination (p = 0.0001), with 78% of individuals exceeding the cut-off, compared to 37% before the booster dose (Figure 1D). PWH showed a similar response trend (Figure 1, red dots), although the small sample size prevented a powered analysis.

**Figure 1:**
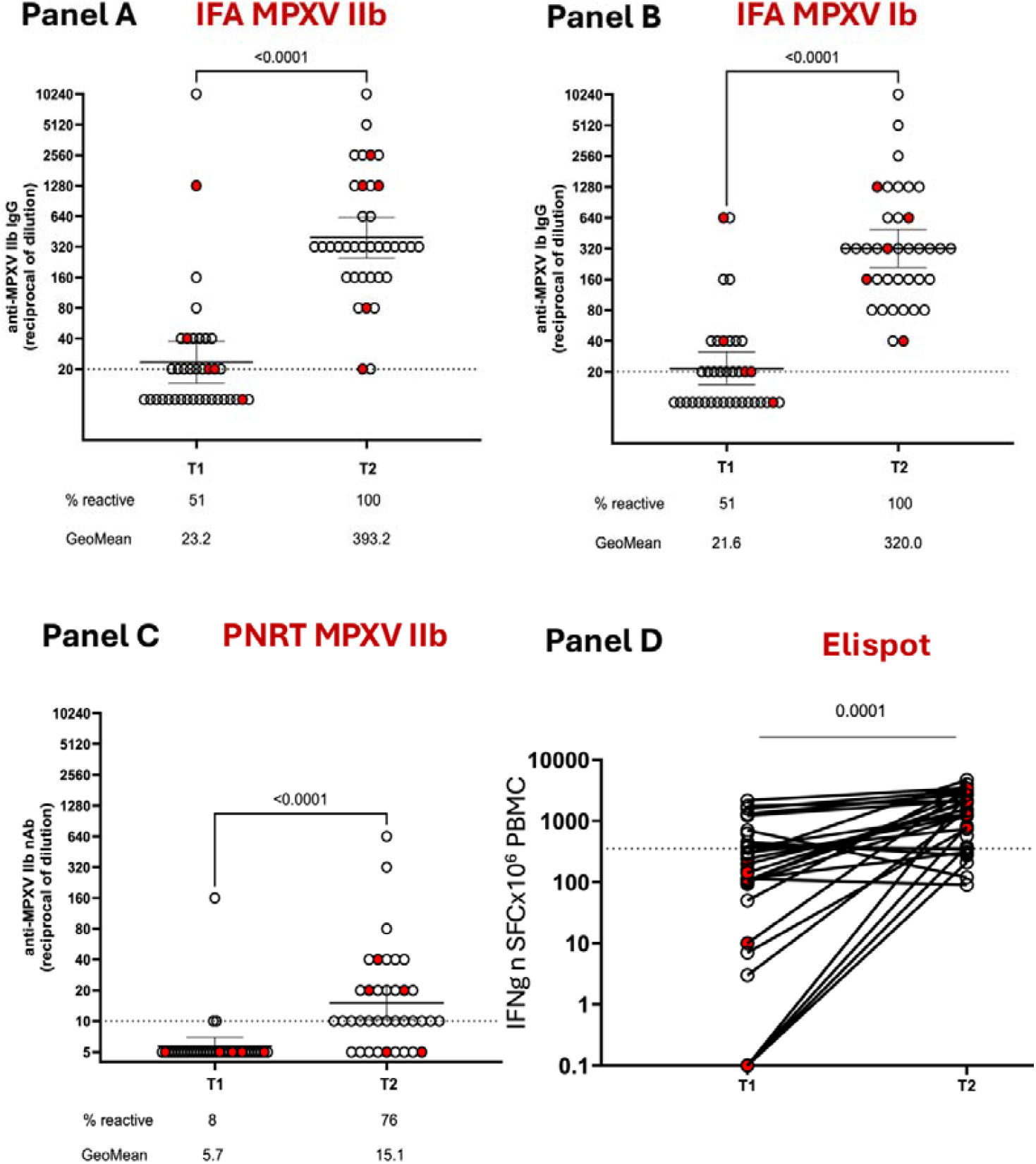
Immune response after an MVA-BN booster dose. MPXV lgG titers against clade IIb (panel A) and Ib (panel B), MPXV neutralising antibodies (nAbs) titers (clade IIb) (panel C) and MVA-BN T-cell specific response expressed as number of Spot Forming Cells (SFC)x10^6^ PBMC (panel D). Red spots= PWH (people living with HIV). Antibody titers are expressed as Geometric Mean Titres (GMT) of the reciprocal serum dilution. Dashed line: limit of assay detection (1:20 for lgG, 1:10 for nAbs and a home-made cut-off based on results from 10 healthy donors for T-cell response). Error bars refer to the 95% confidence interval of GMT. T1=baseline, T2=1 month after booster

### Discussion

Our study demonstrates that an MVA-BN booster administered two years after the primary vaccination series effectively restores and enhances both humoral and cellular immunity in individuals at ongoing risk for mpox. We observed a significant increase in IgG titres against both circulating MPXV clades, suggesting potential cross-clade protection, alongside marked improvements in neutralising antibody levels and T-cell responses.

These findings are relevant in the context of MPXV still circulating^12 13^ and of the debate on booster recommendations. The US CDC currently limits booster administration to individuals with persistent high-risk exposures, reflecting uncertainties regarding the duration of protection and lack of defined correlates of immunity^8^.In contrast, in other countries (e.g. France), health authorities endorsed broader booster use, citing the important waning of nAbs over time, and concerns over the emergence of Clade Ib and its potential clinical and epidemiological implications^9^.

Our findings may help reconcile these divergent positions. We previously demonstrated that, after the primary MVA-BN vaccination, humoral immunity, particularly neutralising immunity, wanes over time, with less than one-third of vaccinees maintaining detectable neutralising antibodies one year after vaccination^3^ and a poor residual response two years after. Conversely, cellular immunity appears more durable despite being substantially diminished two years after vaccination^4^.

In this context, our results showed that a booster administered two years after the primary course significantly enhances both arms of the immune response. Notably, we observed increased IgG titres (reaching 100% seroreactivity) against both clades (IIb and Ib), supporting cross-protection and the usefulness of MVA-BN also against Clade Ib.

The question of booster needs in specific subgroups, such as PWH, remains poorly clarified. While MVA-BN is generally safe and immunogenic in PWH^14 15^, more recent data suggest a suboptimal neutralising response early after a primary single-dose cycle, supporting the recommendation for a full two-dose regimen regardless of prior smallpox vaccination history^16^.

In our study, the immunological response trend after the booster dose seemed similar in PWH and people without HIV (PWoH), although the small sample size prevents definitive conclusions. To date, no validated immunological correlates of protection against mpox have yet been identified, making it challenging to translate laboratory findings into clear policy decisions, and data on efficacy are mainly derived through emulation trials from observational data^17^. In this uncertain landscape, immunogenicity data are crucial for supporting evidence-based vaccination strategies.

Limitations of our study include the small sample size and the lack of a control group of unboosted individuals, which precludes direct assessment of clinical protection or waning in the absence of boosting. The short follow-up period limits conclusions on the durability of the booster-elicited immune response. Finally, the study was not powered to detect subgroup differences, particularly among PWH.

Nonetheless, these data provide essential immunological evidence supporting booster dose administration in individuals at ongoing risk for mpox, contributing to the refinement of vaccination policies in the evolving epidemiological scenario.

## Data Availability

All data produced in the present study are available upon reasonable request to the authors

## SUPPLEMENTS

### Laboratory methods

Anti-MPXV IgG in serum were titered using an indirect immunofluorescence assay (IFA) based on in-house prepared slides with Vero E6 cells infected with either clade IIb MPXV isolated from the skin lesion of a mpox patient hospitalised during the 2022 outbreak (GenBank: ON745215.1) or a clade Ib MPXV obtained from WHO BioHub System (hMpxV/DRCINRB/22MPX0422C/2023; WHO Catalog BMEPP registration number: 2023-WHO-LS-008). The starting dilution for the testing of serum samples was 1:20. The secondary antibody was purchased from Euroimmun.

For measuring anti-MPXV nAb, serum samples were heat-inactivated at 56°C for 30□min and titrated in duplicate in 4 four-fold serial dilutions (starting dilution 1:10). Each serum dilution was added to 100 TCID50 MPXV isolate (GenBank: ON745215.1) and incubated at 37°C for 2□h. Subsequently, 96-well tissue culture plates with Vero E6 cell monolayers were infected with 120□μL/well of virus/serum mixtures and incubated at 37°C and 5% CO2. After 5 days, the supernatant was discarded, a staining-fixing solution (crystal violet containing 10% formaldehyde Diapath S.P.A. and Sigma-Aldrich) added for 30□min, removed, and washed with PBS 1X (Sigma-Aldrich). The number of plaques was counted using the Cytation 5 reader (Biotek), and when the average number of plaques was lower than half the number counted in the virus alone, the serum sample was considered neutralising. The highest serum dilution showing at least 50% of the plaques number reduction was indicated as the 50% neutralization titer (PRNT50%). Each test included serum control (1:10 dilution of each sample tested without virus), cell control (Vero E6 cells alone), and virus control (100 TCID50 MPXV in sextuplicate).

Peripheral blood mononuclear cells (PBMC) were isolated from enrolled subjects using Ficoll density gradient centrifugation (Pancoll human, PAN Biotech) and frozen in FBS (Fetal bovine serum, Euroclone, Italy) added of 10% DMSO (Merck Life sciences, Milan, Italy) at vapors of liquid nitrogen for the experimental design.

Interferon-γ producing T-cell specific response to the MVA-BN vaccine was analyzed by ELISpot assay. PBMC were thawed and suspended in complete medium [RPMI-1640 added of 10% fetal bovine serum, 1% L-glutamine, and 1% penicillin/streptomycin (Euroclone, Italy)]. Live PBMC were counted by Trypan blue exclusion and plated at 3×10^5^ cells/well in ELISpot plates (Human IFN-y ELISpot plus kit; Mabtech, Sweden). MVA-BN vaccine suspension (MOI 1) [JYNNEOS (Smallpox and Monkeypox Vaccine, Live, non-replicating)] added of αCD28/αCD49d (1 µg/ml, BD Biosciences) was used for 20h stimulation at 37 °C (5% CO_2_). After the incubation, the ELISpot assay was developed following manufacturer’s instructions. The spontaneous IFN-γ release was calculated in unstimulated culture (background) and a superantigen (SEB, 200nM, Sigma) was used as positive control. Results are expressed as spot-forming cells per 10^6^ PBMC (SFC/10^6^ PBMC).

## Authors’ contribution

VM and AA conceived the study protocol; VM wrote the first draft of the manuscript; AA, GMa, EC, EG, AS and FM revised the manuscript. JP and AC were responsible for data management.

VM, RE, AG, and GMi enrolled and followed the patients across time. GMa, FC, AB, LB, EC, SN, ET and SG provided laboratory tests. All authors approved the final version of the manuscript.

## Acknowledgements

We thank the study participants and the technical and nursing staff.

